# Skin Pigmentation and Pulse Oximeter Accuracy in the Intensive Care Unit: a Pilot Prospective Study

**DOI:** 10.1101/2023.11.16.23298645

**Authors:** Ashraf Fawzy, Harith Ali, Peter H. Dziedzic, Niteesh Potu, Eusebia Calvillo, Sherita H. Golden, Theodore J. Iwashyna, Jose I. Suarez, David N. Hager, Brian T. Garibaldi

## Abstract

**Rationale:** Despite multiple reports of pulse oximeter inaccuracy among hospitalized Black individuals, regulatory testing of pulse oximeters is performed on healthy volunteers.

**Objective:** Evaluate pulse oximeter accuracy among intensive care unit patients with diverse skin pigmentation.

**Methods:** Skin pigmentation was measured using a chromameter in 12 patients and individual typology angle (ITA), a measure of constitutive pigmentation, calculated. Arterial blood gas (ABG) arterial oxygen saturation (SaO_2_) sampling was precisely matched to pulse oximetry (SpO_2_) using arterial line waveforms analysis. Error (SpO_2_-SaO_2_), bias, and average root mean square error (A_RMS_) were calculated. Multivariable linear mixed effects models evaluated the association of SpO_2_-SaO_2_ with skin pigmentation.

**Measurements and Main Results:** Sampling time was determined for 350 ABGs. Five participants (N=96 ABGs) were darkly pigmented (forehead ITA<-30°), and 7 lighter pigmented (N=254 ABGs). Darkly pigmented individuals had 1.05% bias and 4.15% A_RMS_ compared to 0.34% bias and 1.97% A_RMS_ among lighter pigmented individuals. After adjusting for SaO_2_, pH, heart rate, and mean arterial pressure, SpO_2_-SaO_2_ was falsely elevated by 1.00% more among darkly pigmented individuals (95% confidence interval: 0.25-1.76%). SpO_2_ significantly overestimated SaO_2_ for dark, brown, and tan forehead or forearm pigmentation and brown and tan finger pad pigmentation compared to intermediate/light pigmentation.

**Conclusions:** The pulse oximeter in clinical use at an academic medical center performed worse in darkly pigmented critically ill patients than established criteria for FDA clearance. Pulse oximeter testing in ICU settings is feasible, and could be required by regulators to ensure equivalent device performance by skin pigmentation among patients.

## Introduction

The US Food and Drug Administration (FDA) currently requires medical-grade pulse oximeters be tested on at least 10 unique individuals, with a suggestion that the test sample should include at least 2 darkly pigmented subjects (or 15% of the subject pool, whichever is larger).(1) These individuals need to contribute 200 or more paired oxyhemoglobin saturation measurements from the pulse oximeter (SpO_2_) and an arterial blood sample (SaO_2_). The FDA clearance threshold is an average root mean square error (A_RMS_) ≤3.0% for transmittance, wrap, or finger clip oximeters or ≤3.5% for ear clip or reflectance oximeters. Existing FDA regulatory submissions have exclusively used desaturation testing in healthy volunteers in laboratory settings.(2, 3) Pulse oximeters are intended for clinical use in individuals experiencing acute and/or chronic illness.

The accuracy of pulse oximeters has been questioned,(4) with recurring concern for poor performance among individuals who are darkly pigmented.(5–8) Recent studies showed that pulse oximeters commonly overestimate SaO_2_ among Black and Hispanic patients leading to unrecognized or “occult” hypoxemia.(9–12) Unrecognized hypoxemia is associated with delay in delivering COVID-19 therapy and higher readmission rate among COVID-19-infected patients.(13, 14)

However, past clinical studies have major limitations. First, race and ethnicity as recorded in the electronic medical record (EMR) was used as a surrogate for skin pigmentation, the presumed relevant physiologic parameter. Second, manually entered arterial blood gas (ABG) results and corresponding SpO_2_ readings are obtained from the EMR, with paired SpO_2_-SaO_2_ values occurring up to 10 minutes apart.(15) Some have argued this creates uncertainty if whether SpO_2_ readings that are paired to SaO_2_ measurements accurately reflect the same physiologic state given ongoing care.(16)

This prospective study addresses the limitations of prior retrospective studies by objectively measuring skin pigmentation and identifying pulse oximeter readings at the exact time of ABG collection to evaluate whether pulse oximeter accuracy differs by skin pigmentation in a sample of critically ill patients hospitalized at a single center. The sample was chosen to be larger than that required for FDA premarket notification submission.(1)

## Methods

### Subjects and Measurements

Patients in the Johns Hopkins Hospital (JHH) adult medical intensive care unit (ICU) who had an arterial catheter were prospectively recruited. The Johns Hopkins University Institutional Review Board approved this study and informed consent was obtained before data collection (IRB#00318076). Skin pigmentation was measured at the time of study enrollment according to the Commission Internationale de l’Eclairage (CIE) L*a*b* coordinates (L*=black/white, a*=red/green, b*=yellow/blue) using the Konica Minolta CR-400 chromameter (Ramsey, NJ) at the forehead, forearm, and finger pad.(17) Three consecutive measurements were taken at each site and averaged. Left and right finger pad values were combined for each individual by averaging the values from both hands.

Concurrent SaO_2_ and SpO_2_ were gathered from patients’ entire hospitalization. ABG samples were analyzed on an ABL825 or ABL827 blood gas analyzer (Radiometer America, Brea, CA) and functional SaO_2_ determined by co-oximetry. Pulse oximetry was measured using the Masimo SET LNCS Neo-3 wrap sensor (Irvine, CA) connected to a GE Carescape B850 monitor (Chicago, IL) with E-MASIMO-00 Module. The Masimo LNCS TC-I reusable ear clip sensor was also available but intended only for short-term monitoring.

Vital sign data, including SpO_2_, was recorded at 2 second intervals, and waveforms for both the arterial line and pulse oximeter (photoplethysmography) were visually inspected in the Sickbay^TM^ Platform (Houston, TX). The period of ABG sampling was determined by a physician (A.F. or H.A.) through visual identification of a dampened arterial line waveform proximate to EMR-recorded ABG collection time (**Supplementary Figure 1**). Readers were blinded to participant race and skin pigmentation. Photoplethysmography waveform amplitude was subjectively assessed as ‘poor’, ‘variable’, or ‘good’ (**Supplementary Figure 2**).

For each ABG, the corresponding SpO_2_ value was the median SpO_2_ during ABG sampling as determined by dampening of the arterial waveform. The median heart rate from pulse oximetry was determined during ABG sampling and median mean arterial pressure was determined from the arterial line 60-120 seconds before or after ABG sampling. Occult hypoxemia was defined as SaO_2_<88% with concurrent SpO_2_≥92%.

### Statistical Analysis

The individual typology angle (ITA), a measure of constitutive skin pigmentation,(18) was calculated from the L* and b* CIE coordinates as [arctan(L*-50)/b*)]*(180/π) separately for each participant and anatomic site.(17) Participants were categorized for the primary analysis based on forehead ITA as darkly pigmented if calculated ITA was less than -30° and lighter pigmented otherwise.(17)

Error was calculated as the difference between SpO_2_ and SaO_2_ (SpO_2_-SaO_2_). The bias (mean difference; √(SpO_2_-SaO_2_)/N) and A_RMS_ (√([SpO_2_-SaO_2_]^2^)/N) were calculated according to the International Organization for Standardization (ISO) document 80601-2-61:2011.(1, 19) Bootstrapping with 10,000 sampling replications for each skin pigmentation category was used for statistical comparison of the A_RMS_.(20) Separate linear mixed effects models were constructed for SpO_2_-SaO_2_ with skin pigmentation category or forehead, forearm, or finger pad ITA as the independent variable accounting for repeated measures within subject. Non-linear associations were tested retaining quadratic and cubic terms when statistically significant (p<0.05). Multivariable models controlled for SaO_2_, pH, heart rate, and mean arterial pressure. Marginal effects from transition of intermediate/light pigmentation (ITA=41°) to the midpoint of tan (ITA=19°), brown (ITA=-10°) and dark (ITA=-45°) pigmentation were calculated.

A secondary analysis excluded lower quality SpO_2_ values defined as SpO_2_-SaO_2_ pairs where the range (defined as maximum-minimum) of SpO_2_ was >1% for 30 seconds or the duration of blood sampling, whichever was longer, or where the photoplethysmography tracing was categorized as ‘poor’. An alternative analysis was performed using the closest SpO_2_ value within 10 seconds of the time SaO_2_ was recorded in the EMR. Analyses were conducted using SAS 9.4 (Carey, NC).

## Results

Twelve individuals were enrolled during their ICU stay from June to October 2022. Five were darkly pigmented and seven lighter pigmented based on forehead pigmentation measurement. Darkly pigmented individuals had a median age of 54 years (range: 42.9-70.2), 2 (40%) were female, and all were identified as non-Hispanic Black in the EMR. Lighter pigmented individuals had a median age of 64.8 (range: 34.7-79.4), 4 (57.1%) were female, and one was identified as Asian whereas the remainder were identified as non-Hispanic White in the EMR. Median forehead ITA was -46.2° for darkly pigmented individuals (range: -52.5° to -42.0°) and 12.6° for lighter pigmented individuals (range: -6.3° to 48.3°). The distribution of pigmentation was similar at the forearm but differed at the finger pad (**Figure 1**).

**Figure 1:**
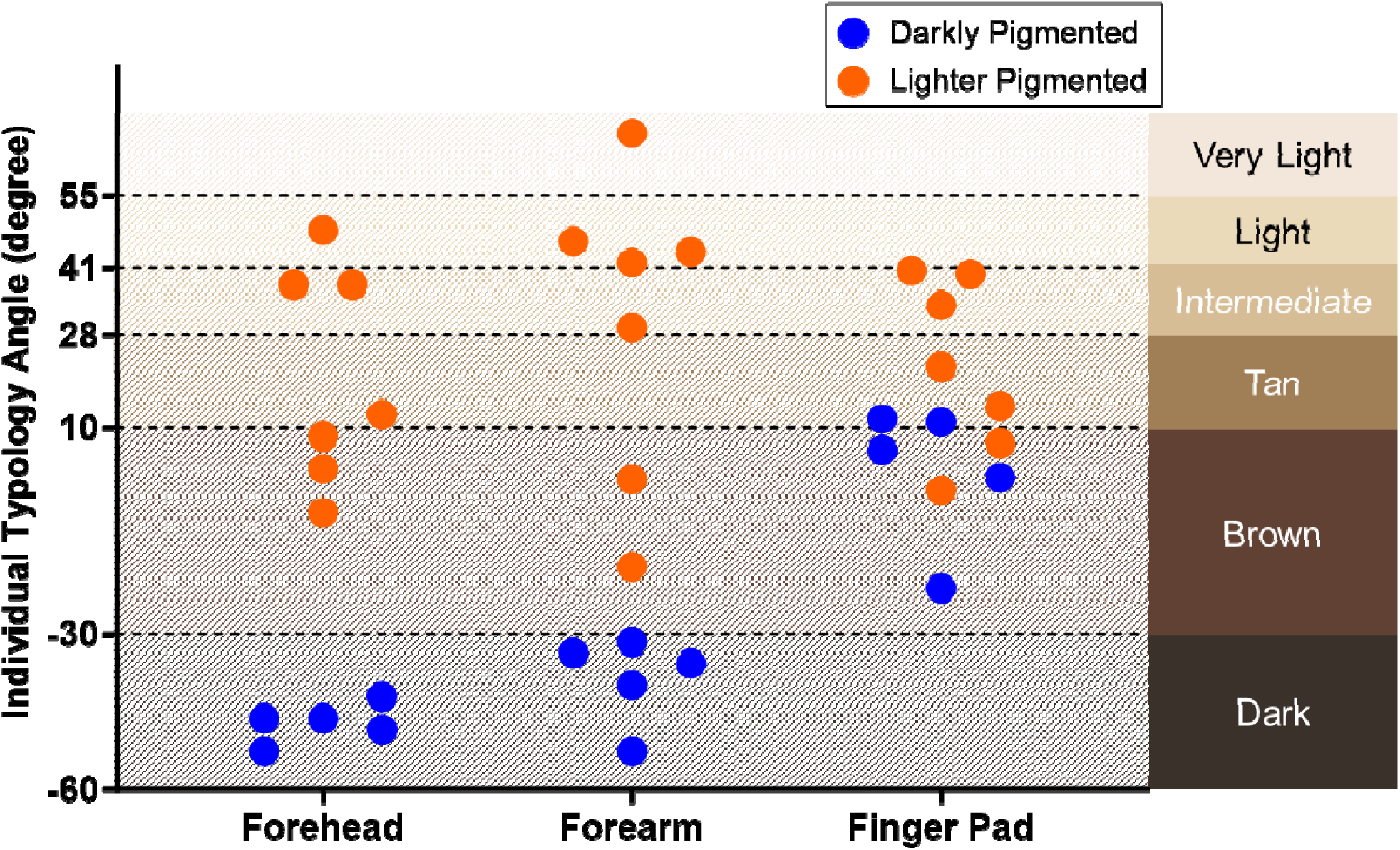
Distribution of objectively measured skin pigmentation at the forehead, forearm, and finger pad of participants.

Four hundred ABGs were collected during routine clinical care (119 ABGs for darkly pigmented individuals and 281 for lighter pigmented individuals) with a median of 24 ABGs for darkly pigmented individuals (range: 2-49) and 12 ABGs for lighter pigmented individuals (range: 1-129). Fifty observations were excluded from the primary analysis when a corresponding SpO_2_ could not be identified because at the time of ABG the participant was not in an ICU, an SpO_2_ value was not available, or the arterial line tracing was absent/non-pulsatile. There were 350 ABGs (96 in darkly pigmented participants, 254 in lighter pigmented participants) where the blood sampling time could be identified and corresponding SpO_2_ measurements were available. Notably, the median duration between the ABG time recorded in the EMR and the exact time of ABG sampling obtained from assessing the arterial line tracing was 77.5 seconds (interquartile range: 39.6-123.8 seconds), ranging to over 10 minutes (618.2 seconds) away. SpO_2_<90% was more common among darkly pigmented individuals than lighter pigmented individuals (5.2% vs 1.6%) and poor plethysmography quality was rare (**Table 1**).

**Table 1:** Characteristics of arterial blood gas samples and paired pulse oximetry by skin pigmentation.

|  | All Patients | Darkly Pigmented Patients | Lighter Pigmented Patients |
| --- | --- | --- | --- |
| Total ABG Samples Recorded | 400 | 119 | 281 |
| SaO<sub>2</sub> (median, IQR) | 97.2 (95.0-98.9) | 97.2 (95.0-98.9) | 97.2 (95.4-98.5) |
| SaO<sub>2</sub> < 90% | 11 (2.8%) | 6 (5%) | 5 (1.8%) |
| SaO<sub>2</sub> 90-94% | 45 (11.3%) | 12 (10.1%) | 33 (11.7%) |
| SaO<sub>2</sub> > 94% | 344 (86%) | 101 (84.9%) | 243 (86.5%) |
| pH (median, IQR) | 7.42 (7.38-7.47) | 7.40 (7.26-7.45) | 7.43 (7.39-7.47) |
| Blood Sampling Period Identified (N) | 350 | 96 | 254 |
| Blood Sampling Duration, seconds (median, IQR) | 46.1 (28.1-69) | 43.6 (28.7-65.8) | 46.7 (28-70.3) |
| SaO<sub>2</sub> (median, IQR) | 97.1 (95.1-98.4) | 97.0 (94.9-98.7) | 97.2 (95.3-98.3) |
| SaO<sub>2</sub> < 90% | 9 (2.6%) | 5 (5.2%) | 4 (1.6%) |
| SaO<sub>2</sub> 90-94% | 44 (12.6%) | 11 (11.5%) | 33 (13%) |
| SaO<sub>2</sub> > 94% | 297 (84.9%) | 80 (83.3%) | 217 (85.4%) |
| pH (median, IQR) | 7.43 (7.39-7.47) | 7.43 (7.31-7.45) | 7.43 (7.40-7.48) |
| Heart Rate, beats per minute (median, IQR) | 90 (74-105) | 98 (79-111.5) | 89 (72-101.5) |
| Mean Arterial Pressure, mmHg (median, IQR) | 78 (70-91) | 73.8 (70-81) | 80.3 (70-96) |
| Photoplethysmography Quality |  |  |  |
| Good | 137 (39.1%) | 48 (50%) | 89 (35%) |
| Variable | 207 (59.1%) | 45 (46.9%) | 162 (63.8%) |
| Poor | 6 (1.7%) | 3 (3.1%) | 3 (1.2%) |
| SpO<sub>2</sub> maximum-minimum* (median, IQR) | 1 (0-1) | 1 (0-1) | 1 (0-1) |
| 0% | 139 (39.7%) | 38 (39.6%) | 101 (39.8%) |
| 1% | 139 (39.7%) | 36 (37.5%) | 103 (40.6%) |
| ≥2% | 72 (20.6%) | 22 (22.9%) | 50 (19.7%) |
| Blood Sampling Occurrence |  |  |  |
| Before Recorded SaO<sub>2</sub> Time | 276 (78.9%) | 77 (80.2%) | 199 (78.4%) |
| During Recorded SaO<sub>2</sub> Time | 22 (6.3%) | 4 (4.2%) | 18 (7.1%) |
| After Recorded SaO<sub>2</sub> Time | 52 (14.9%) | 15 (15.6%) | 37 (14.6%) |
| Duration between blood sampling and recorded SaO<sub>2</sub> time, seconds (median, IQR) | 77.5 (39.6-123.8) | 81.3 (39.1-122.7)<br>Maximum: 571.3 | 74.1 (39.9-124.9)<br>Maximum: 618.2 |
| Occult Hypoxemia (SaO<sub>2</sub><88%, SpO<sub>2</sub>≥92%) | 3 (0.9%) | 2 (2.1%) | 1 (0.4%) |
| Pulse Oximetry within 10 seconds of recorded time | 360 | 100 | 260 |

| (N) |  |  |  |
| --- | --- | --- | --- |
| Duration between SpO<sub>2</sub> and recorded SaO<sub>2</sub> time, seconds (median, IQR) | 0 (0-1) | 0 (0-1) | 0 (0-1) |
| SaO<sub>2</sub> (median, IQR) | 97.1 (95.1-98.4) | 97.0 (95.0-98.5) | 97.1 (95.2-98.3) |
| SaO<sub>2</sub> < 90% | 10 (2.8%) | 5 (5%) | 5 (1.9%) |
| SaO<sub>2</sub> 90-94% | 44 (12.2%) | 11 (11%) | 33 (12.7%) |
| SaO<sub>2</sub> > 94% | 306 (85%) | 84 (84%) | 222 (85.4%) |
| pH (median, IQR) | 7.43 (7.39-7.47) | 7.43 (7.30-7.45) | 7.43 (7.40-7.48) |
| Occult Hypoxemia (SaO<sub>2</sub><88%, SpO<sub>2</sub>≥92%) | 2 (0.6%) | 2 (2%) | 0 (0%) |

\* For 30 seconds or duration of blood sampling period, whichever is longer

Overall, pulse oximeter bias was 0.62% and the A_RMS_ was 2.75%. However, among darkly pigmented individuals pulse oximeter bias was 1.05% and the A_RMS_ was 4.15%— substantially higher than for lighter pigmented individuals (bias 0.34% and A_RMS_ 1.97%). The 95% CI for A_RMS_ from 10,000 bootstrap was 2.35-5.72% for darkly pigmented individuals and was significantly higher than 1.76-2.17% for lighter pigmented individuals (p<0.0001). Occult hypoxemia was present in 2.1% of sample pairs of darkly pigmented participants compared with 0.4% of sample pairs in lighter pigmented participants. Pulse oximeter error for darkly pigmented participants increased as SaO_2_ decreased, with median (range, N) SpO_2_-SaO_2_ of 2.5% (0.8%-7.0%, N=11) for SaO_2_ 90-94% and 10.0% (0.2%-14.5%, N=5) for SaO_2_<90%, unlike in lighter pigmented participants (**Figure 2**).

**Figure 2:**
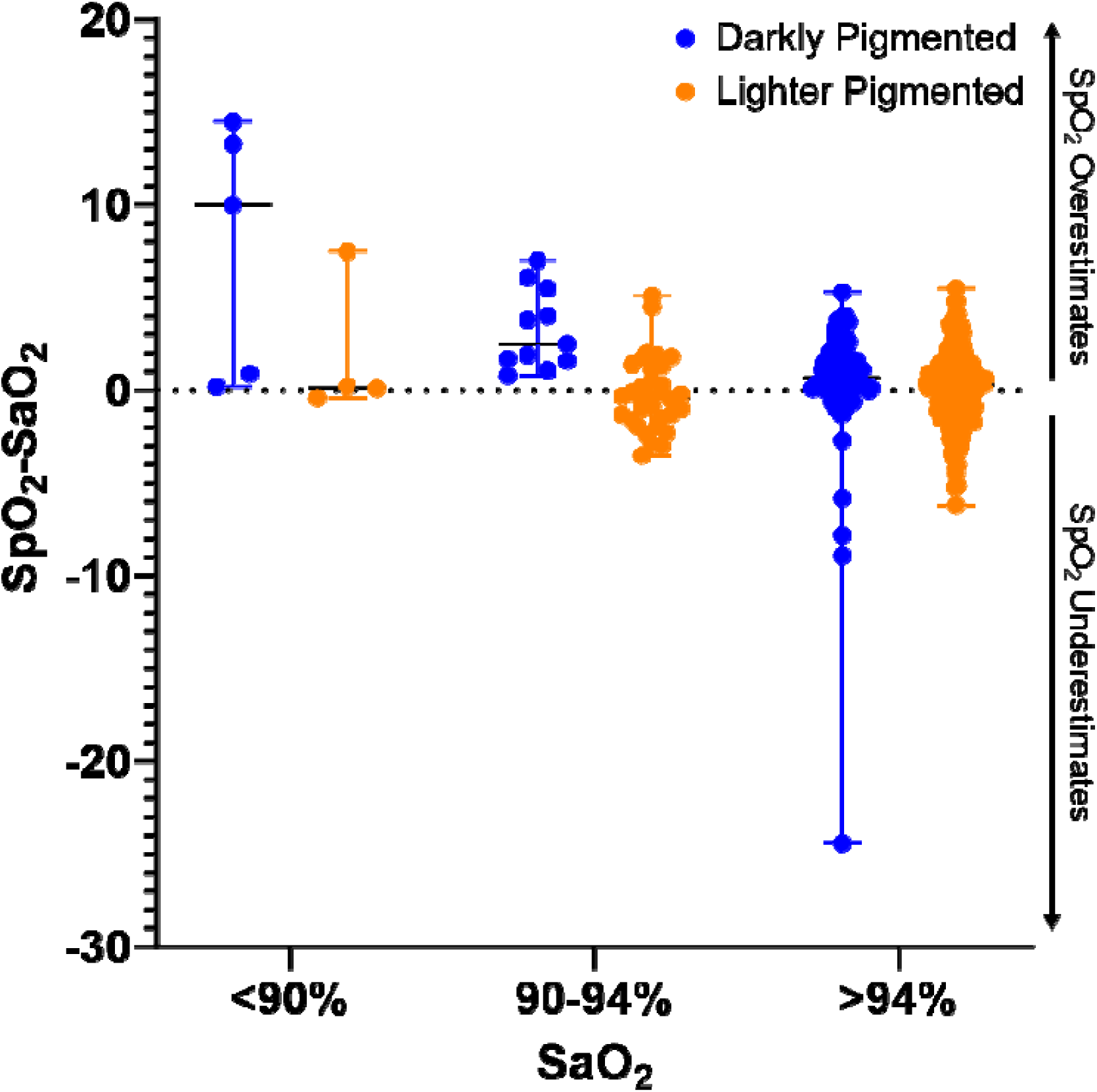
Distribution (with median and range) of pulse oximeter error (SpO_2_-SaO_2_) stratified by skin pigmentation and SaO_2_.

In regression analysis, controlling for SaO_2_, pH, heart rate, and mean arterial pressure, the error of the pulse oximeter (SpO_2_-SaO_2_) was on average 1.00% higher for darkly pigmented individuals compared with lighter pigmented individuals (95% CI: 0.25-1.76%). A 10° reduction in forearm ITA (darker pigmentation) was associated with a 0.19% increase in SpO_2_-SaO_2_ (95% CI: 0.10-0.29%). Associations of pulse oximeter accuracy with forehead and finger pad skin pigmentation were non-linear but demonstrated significant overestimation of SaO_2_ by SpO_2_ for dark, brown, and tan forehead pigmentation and brown and tan finger pad pigmentation (**Figure 3**). The marginal difference compared to intermediate/light pigmentation was 2.40% (95% CI: 1.69-3.12%) for dark, 2.92% (95% CI: 2.11-3.74%) for brown, and 1.80% (95% CI: 1.24-2.35%) for tan forehead pigmentation and 0.99% (95% CI: 0.58-1.40%) for brown and 0.43% (95% CI: 0.25-0.60%) for tan finger pad pigmentation.

**Figure 3:**
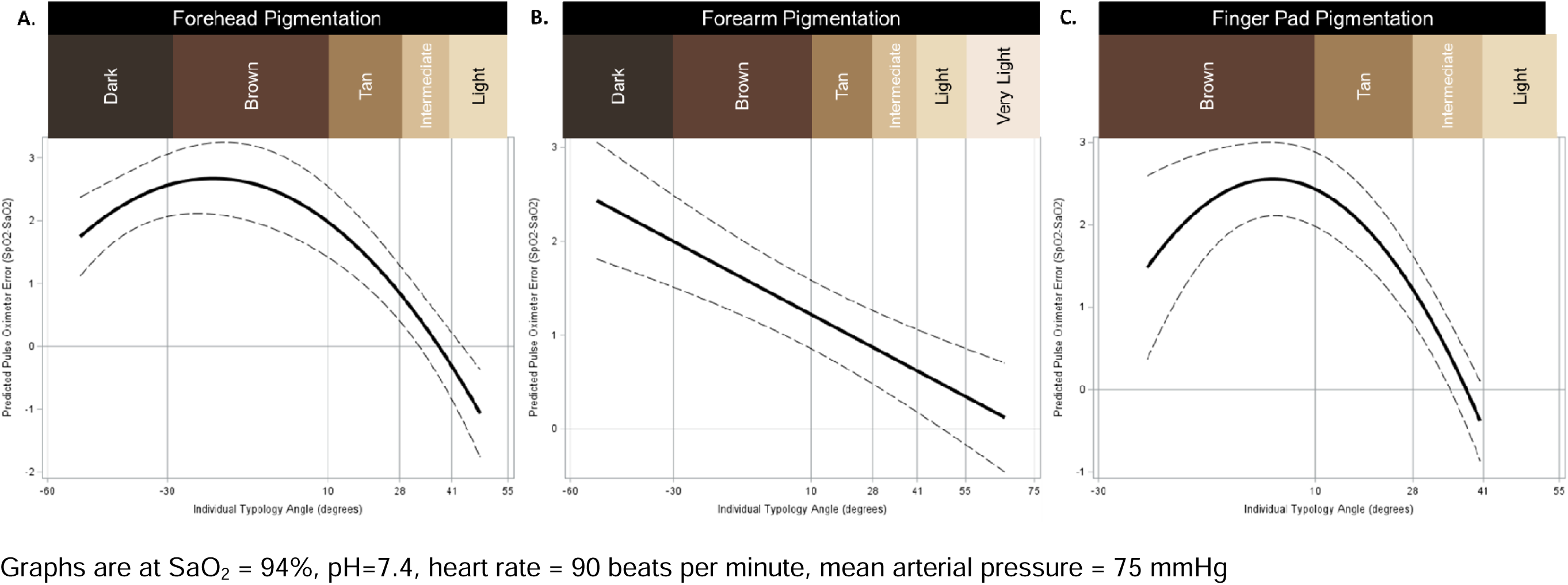
Estimated mean pulse oximeter error (SpO_2_-SaO_2_) by skin pigmentation represented by the individual typology angle (in degrees) at the (a) forehead, (b) forearm, and (c) finger pad.

In sensitivity analyses excluding lower quality SpO_2_ readings (remaining N=73 darkly pigmented and N=203 lighter pigmented), the overall bias and A_RMS_ improved to 0.59% and 2.37% respectively, however the difference in performance between darkly pigmented and lighter pigmented individuals persisted: 1.13% bias and 3.44% A_RMS_ for darkly pigmented compared to 0.08% bias and 1.83% A_RMS_ for lighter pigmented individuals. Multivariable regression results were unchanged (**Supplementary Figure 3**). In the alternative analysis that considered 360 SpO_2_ values within 10 seconds of the recorded SaO_2_ time in the EMR (N=100 darkly pigmented, N=260 lighter pigmented) the overall bias was 0.63% and A_RMS_ was 2.98% with bias and A_RMS_ of 1.05% and 4.59% respectively for darkly pigmented individuals compared to 0.36% and 2.05% for lighter pigmented individuals.

## Discussion

This is the first real-world clinical study using objectively measured skin pigmentation and precise identification of SpO_2_-SaO_2_ pairs to evaluate the accuracy of pulse oximetry in critically ill adult patients with varying skin tone. The FDA-cleared pulse oximeter evaluated in this study performed worse in darkly pigmented individuals and would not meet established criteria for FDA clearance (A_RMS_<3.0%) among critically ill patients with dark skin pigmentation. Conversely, among critically ill patients with lighter skin pigmentation, the pulse oximeter met the FDA’s performance standard. Importantly, increased pulse oximeter error among darkly pigmented individuals was independent of underlying saturation level, acidosis, heart rate, and blood pressure, all of which may compound pulse oximeter error. The study’s sample size meets the current threshold for premarket notification submission for FDA clearance.

This study substantially improves on prior investigations by (1) objectively measuring skin pigmentation rather than relying on race/ethnicity designations in the EMR, (2) leveraging arterial line waveforms to precisely identify the exact time of ABG sampling, and (3) using high-resolution vital sign data to pair ABGs with concurrent SpO_2_ data. This methodologic approach pairs the vast majority (87.5%) of clinical SaO_2_ measurements with SpO_2_ measurements, unlike some EMR studies.(13) Examining arterial waveform tracings also revealed that the recorded SaO_2_ time in the EMR was often a minute before or after the actual time of blood sampling, with a quarter of samples over 2 minutes away. Taken together, the inaccuracy of recorded SaO_2_ time and window for SpO_2_ could have acted as compounding biases in past studies, albeit likely towards underestimating racial disparities.(15) Here, definitively overcoming the potential biases in prior retrospective studies in critically ill patients, the conclusion that pulse oximeters more commonly overestimate oxygen saturation in darkly pigmented individuals is corroborated by this study.

Most recent studies investigating the impact of skin pigmentation on pulse oximeter accuracy among adults in clinical settings have substituted race and ethnicity for objective skin tone measurements.(9–15, 21–23) Laboratory studies of healthy volunteers that subjectively evaluated skin pigmentation have reported poor accuracy at low saturations (<80%) among darkly pigmented individuals compared with lighter pigmented individuals.(7, 8) However, a re-analysis of proprietary healthy volunteer laboratory data reported no difference in pulse oximeter accuracy between Black and White volunteers.(2) This is at odds with the findings of this study and a recent pre-print of work of commercial pulse oximeters with objectively measured skin pigmentation.(3) Early studies using subjective skin pigmentation scales noted that pulse oximeters were less accurate and less reliable among darkly pigmented individuals undergoing cardiopulmonary exercise testing or in the ICU.(5, 24) A study of 100 darkly pigmented South African patients in the ICU found acceptable precision and bias for four oximeters, however, only 5 samples had oxygen saturations ≤92%.(25) In the modern era, a small study of hypoxemic infants with cyanotic congenital heart disease noted poor oximeter performance unrelated to visually assessed skin pigmentation,(26) though routine overestimation of oxygen saturation by pulse oximeters in this chronically hypoxic population is common.(27, 28) One study of adult patients in two ICUs in Australia and New Zealand found that darkly pigmentated patients experienced significantly more overestimation by pulse oximetry compared with lighter pigmented patients.(29)

By objectively measuring skin pigmentation using a hand-held chromameter, we avoid introduction of subjective biases when visually assessing skin pigmentation using a skin tone scale.(3) Although skin pigmentation measurements at the forehead and forearm were best at distinguishing between darkly pigmented and lighter pigmented individuals, finger pad pigmentation was also strongly associated with pulse oximeter error despite lighter pigmentation and greater overlap between darkly and lighter pigmented individuals. Capturing finger pigmentation using a visual scale may be more difficult and less accurate, but findings from this study show similar results regardless of where skin pigmentation is assessed; the generalizability of this to other clinical settings needs to be confirmed.

Although this study addresses prior investigations’ methodologic shortcomings, limitations remain. Manually judging exact start and stop times of ABG sampling from arterial line tracings introduces subjectivity — though differences in judgement are likely to be on the order of milliseconds and raters were blinded to the skin pigmentation of the patients whose waveforms were being evaluated, limiting the risk of differential error. The arterial line may also have been accessed for a prolonged period to obtain blood tests in addition to the ABG. Since the ABG sampling is not directly observed, pulse oximeter placement on the participant’s body is unknown, and the wrap sensor could have been placed on a finger, toe, or ear. As this study relies on clinical data, the distribution of oxygen saturation is heavily skewed toward higher values though this tends to be the range where pulse oximeters are most accurate and would have biased results toward the null. Due to the relatively small patient sample size as currently suggested by the FDA, non-pigment patient level factors could not be included in multivariable assessment and effect modification could not be assessed. Finally, this study only evaluated a single pulse oximeter model and may not be generalizable to other FDA-cleared oximeters.

### Policy Implications

Updated guidelines for regulatory clearance of pulse oximeters could ensure that darkly pigmented subjects whose objectively measured skin tone falls within predefined criteria are included with sufficient power to detect differences in device performance within this group. Furthermore, device testing in the real-world setting is imperative as there are substantial physiologic and perfusion differences between healthy volunteers and individuals with acute or chronic illness. This study serves as a framework for larger future studies testing existing and emerging pulse oximeters using methods that objectively quantify skin pigmentation. It also provides a reproducible framework for leveraging a remote vital sign monitoring system and high-resolution data repository to capture hypoxic events in the clinical setting with lower participant and staff burden. In contrast, a prospective study that solely relies on direct observation of clinical ABGs and research ABGs guided by pulse oximeter readings will invariably miss many instances of occult hypoxemia. In addition, the exclusive use of clinical data in this study most closely reflects real-world use of pulse oximeters whereas the presence of research staff and additional scrutiny at the time of ABG sampling could lead to bias.

### Conclusion

In conclusion, using high-resolution clinical data from a diverse sample of intensive care unit patients with objectively measured skin pigmentation, pulse oximeter error was greater among darkly pigmented patients. This performance would not meet the Food and Drug Administration’s current threshold for regulatory clearance in this population. Furthermore, pulse oximeter inaccuracy varied based on degree of skin pigmentation, independent of other physiologic parameters, demonstrating statistically significant overestimation of true oxygen saturation among individuals with dark or brown facial or forearm skin and brown or tan finger pad skin.

## Data Availability

All data produced in the present study are available upon reasonable request to the authors

