## Supplementary Figure for "Skin Pigmentation and Pulse Oximeter Accuracy in the Intensive Care Unit: a Pilot Prospective Study"

**Supplementary Online Materials**

**Table of Contents:**

**Figure E1:** Representative examples of visual identification of arterial blood gas sampling times by visual inspection of arterial line waveform.

**Figure E2:** Representative examples of subjectively assessed photoplethysmography waveform quality

**Figure E3:** Estimated mean pulse oximeter error ($SpO_2$-$SaO_2$) by skin pigmentation represented by the individual typology angle (in degrees) at the (a) forehead, (b) forearm, and (c) finger pad excluding lower quality $SpO_2$ readings

**Figure E1:** Representative examples of visual identification of arterial blood gas sampling times by visual inspection of arterial line waveform.

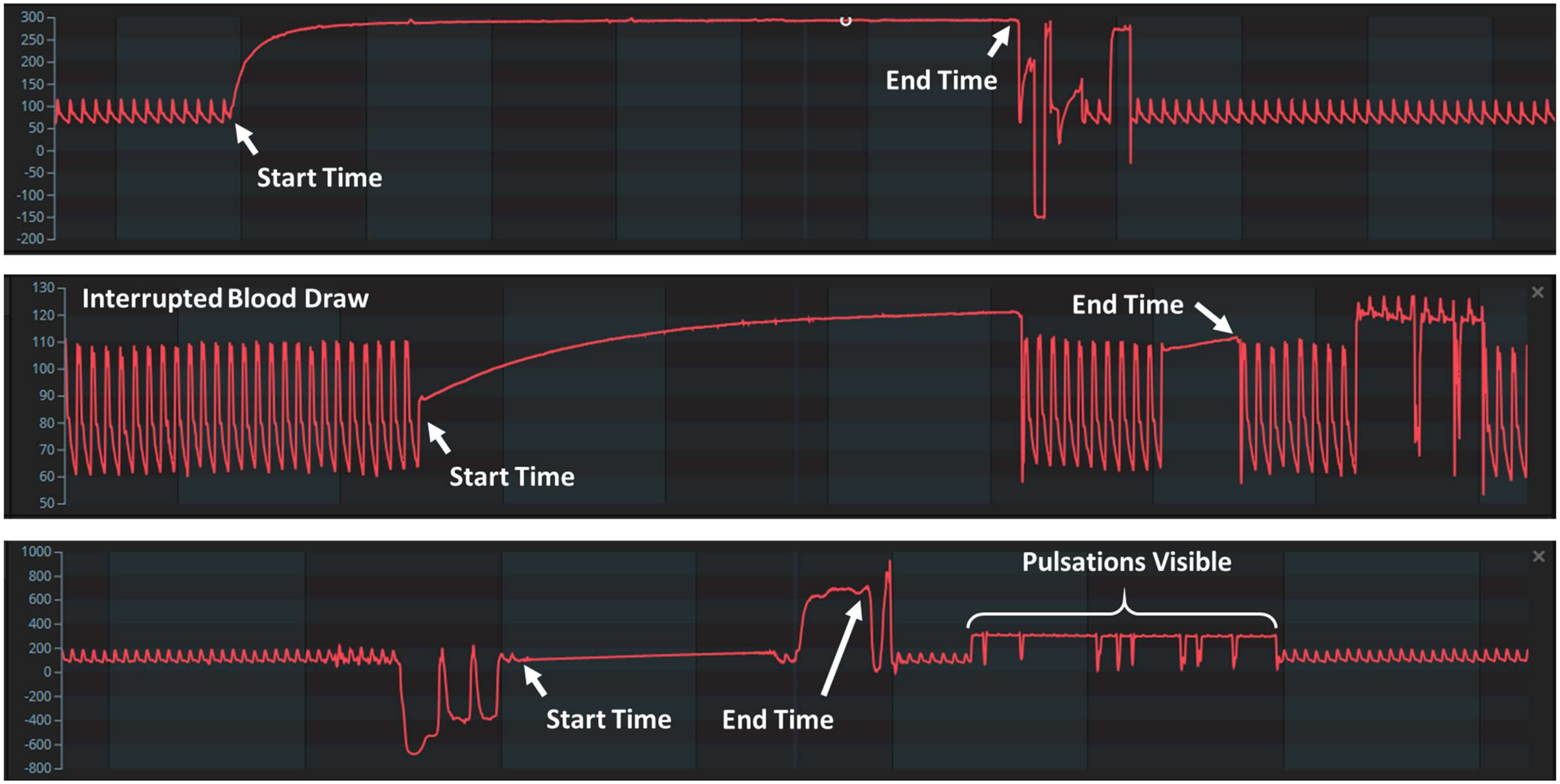

**Figure E2:** Representative examples of subjectively assessed photoplethysmography waveform quality

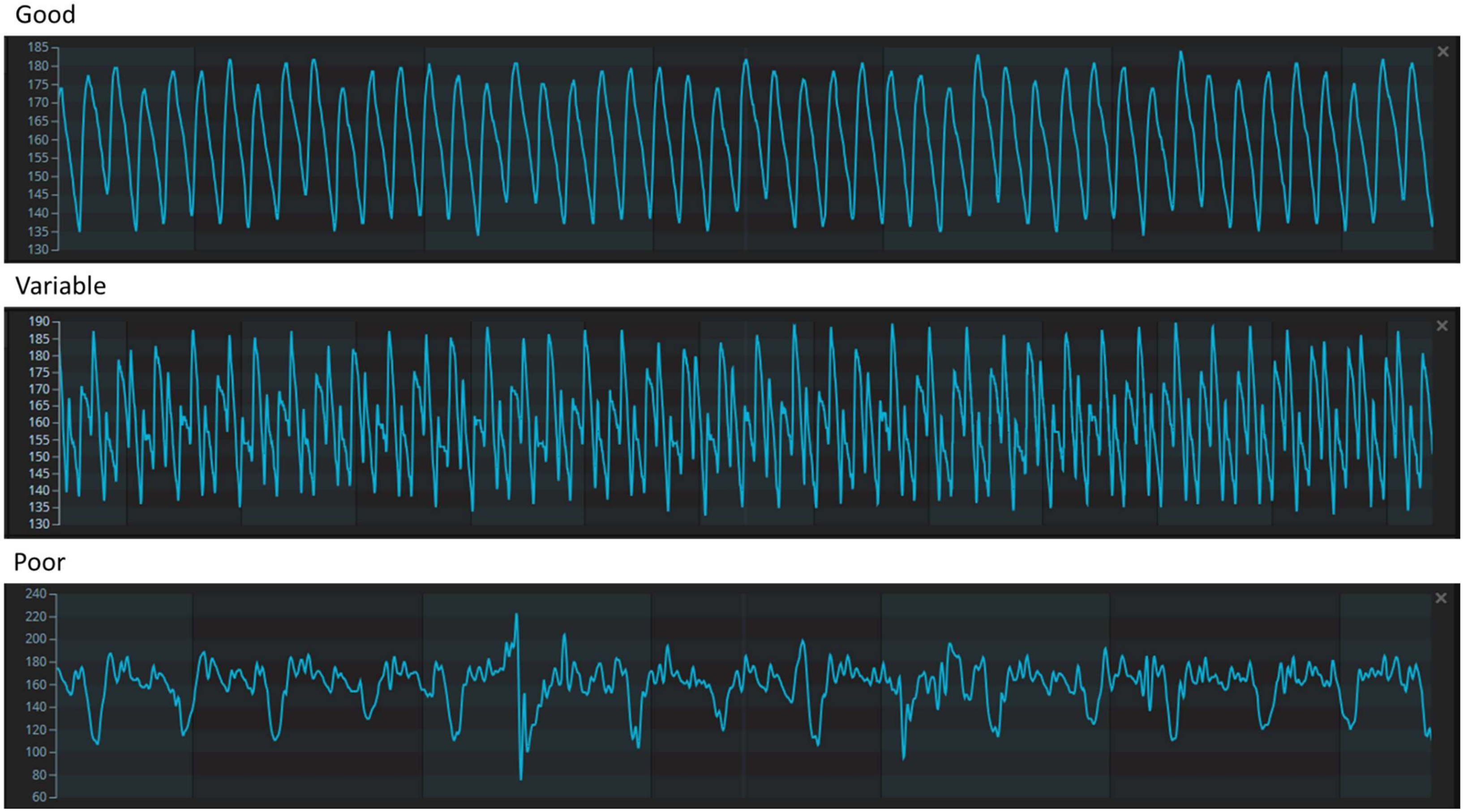

**Figure E3:** Estimated mean pulse oximeter error ($SpO_2$-$SaO_2$) by skin pigmentation represented by the individual typology angle (in degrees) at the (a) forehead, (b) forearm, and (c) finger pad excluding lower quality $SpO_2$ readings

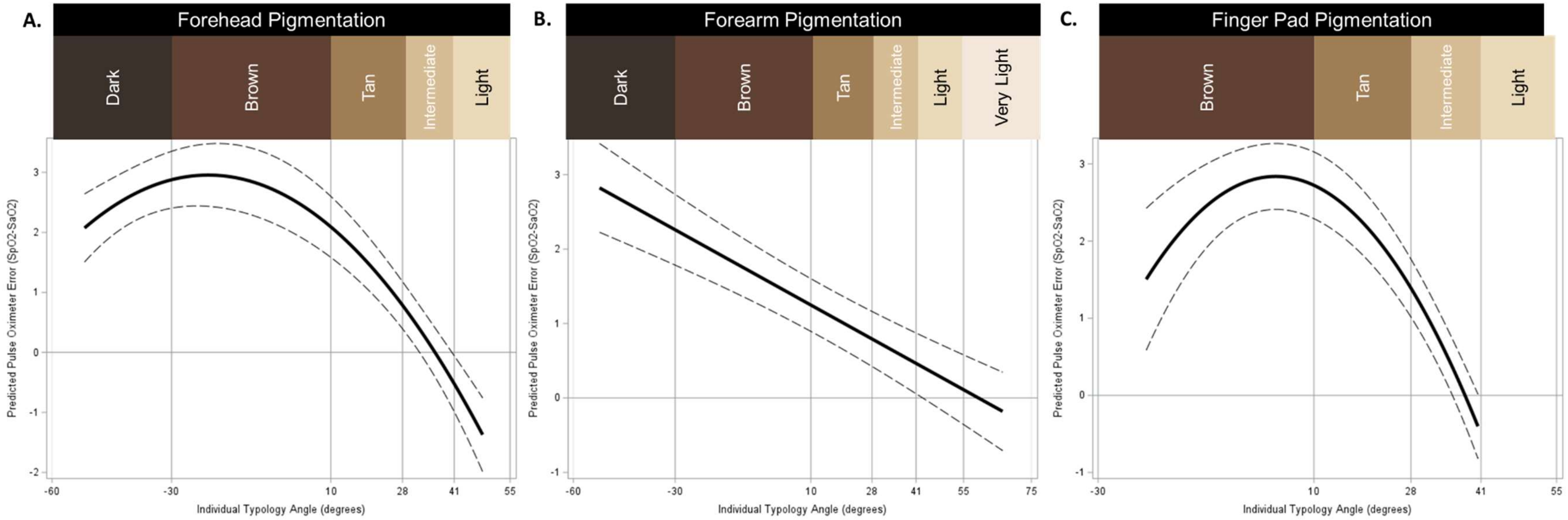


Graphs are at $SaO_2$ = 94%, pH=7.4, heart rate = 90 beats per minute, mean arterial pressure = 75 mmHg
